# A Scaling Law for PCR Positivity in the COVID Second Wave

**DOI:** 10.1101/2021.12.07.21267073

**Authors:** Keith Johnson

**Affiliations:** IP Consultant for Diagnostic Testing, Thurnbichl 2, A-6345 Kössen, Austria

## Abstract

In this preliminary report, PCR positivity data in the second wave of the COVID pandemic (September-January 2020) are shown to obey a scaling law given by:

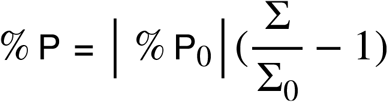

where % P_0_ and Σ_0_ are the y- and x-intercepts of a plot of positivity, %P, against the number of tests, Σ. The law holds across international, regional and local boundaries, as demonstrated for Great Britain, Austria, Germany and Sweden, the nine English regions, London - Yorkshire & Humber, and various Local Health Authorities in England. One possible explanation for scaling might be Dorfman pooling.

The scaling law can be used to remove a systematic or false positive (FP) component from the daily number of positive tests, or cases, to yield the real number of cases. The results correlate strongly with the ZOE survey for London (R^2^=0.787) and Excess Deaths for England (R^2^=0.833). The cumulative total of FPs can be estimated as 1.4M by the beginning of 2021, in line with other estimates.

## I. Introduction

As early as Spring 2020, Levitt and other authors applied a scaling approach to the spread of the SARS-COV2 virus [1-3]. Ohnishi et al [2] showed that the spread in many countries could be characterised by a universal scaling function based on the Gompertz function. The rate of increase of infected people was better characterised by the K parameter, which also showed scaling behaviour, rather than the R parameter of the classical RSI models. Since the number of positive PCR tests has also been used as a proxy for cases, or the number of infected people, especially in the second wave of the pandemic, this raises the question whether scaling behaviour exists here too. As demonstrated below, the answer to this question is yes.

## II. Regression model

The linear correlation between PCR positivity, %P, and the number of tests, Σ, has been noted at international [4], national and regional [5], and local [6] levels. For the UK, Austria, Germany and Sweden, the results are shown in the the following graph:

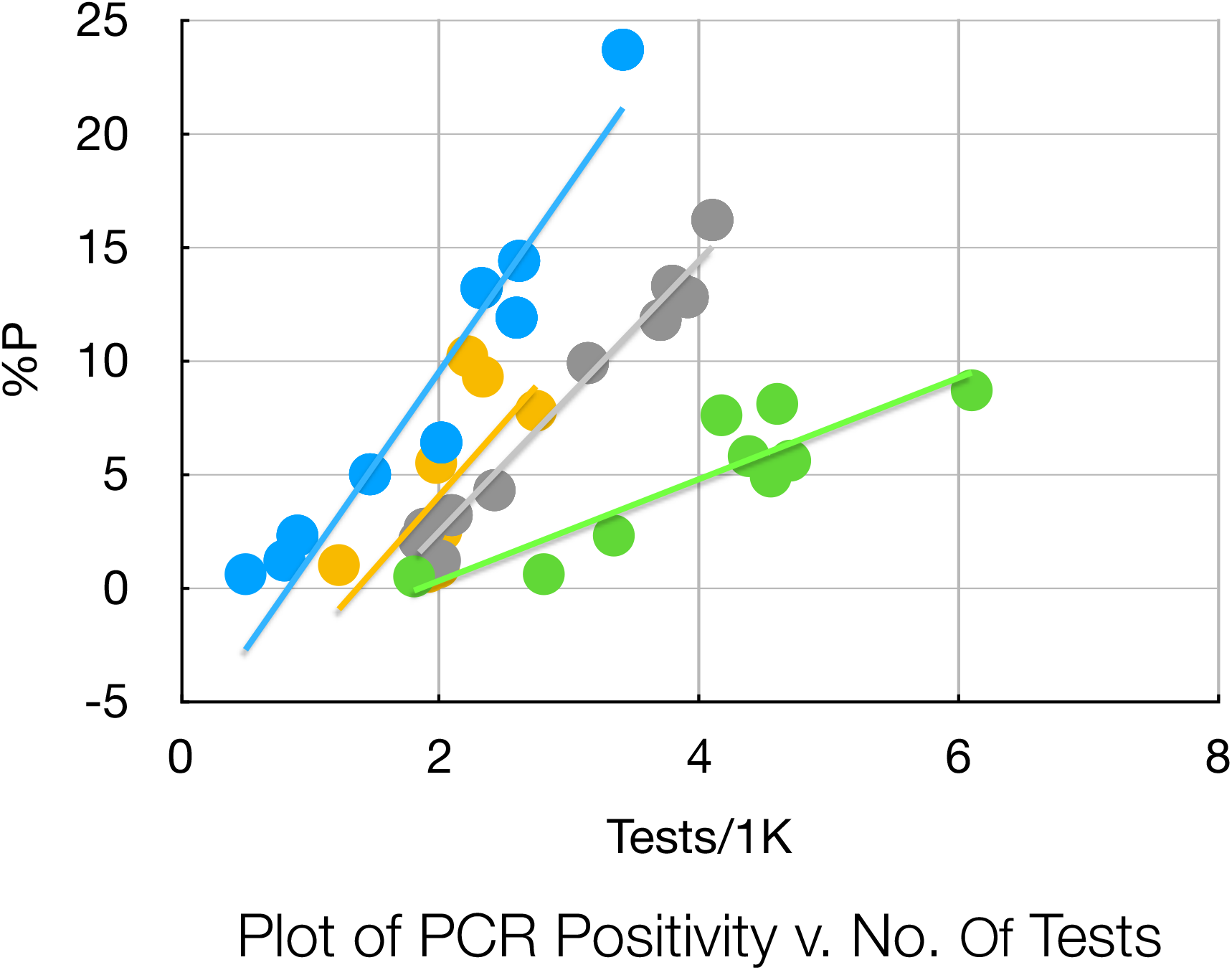

where the raw data were taken from OurWorldinData website, essentially for the months June-December 2020^1^. The straight lines are obtained by least squares regression and obey the following equation:

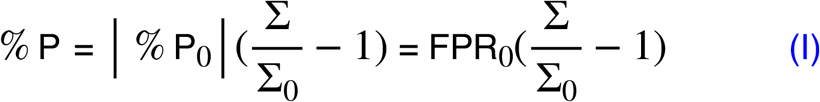

where %P_0_ is the y-intercept and Σ_0_ is the x-intercept. The fit parameters are given in the following table:

**Table I.**
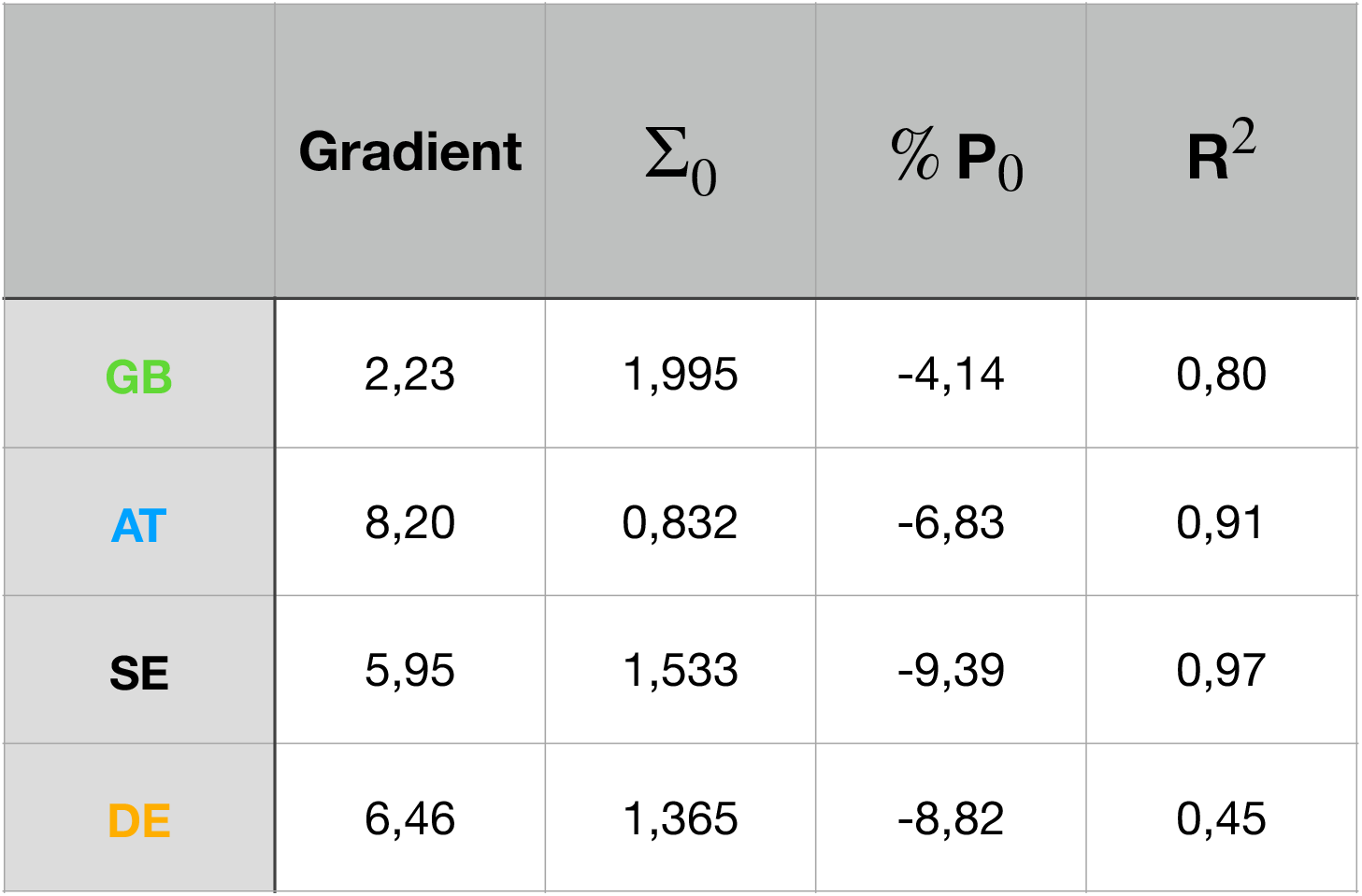

| | Gradient | $\Sigma_0$ | $\% P_0$ | $R^2$ |
| --- | --- | --- | --- | --- |
| GB | 2,23 | 1,995 | -4,14 | 0,80 |
| AT | 8,20 | 0,832 | -6,83 | 0,91 |
| SE | 5,95 | 1,533 | -9,39 | 0,97 |
| DE | 6,46 | 1,365 | -8,82 | 0,45 |

In fact, by scaling the y-axis with |% P_0_| and the x-axis with Σ_0_, all the data can be brought onto a common straight line with a co-efficient of determination, R^2^ = 0,87, whereby:

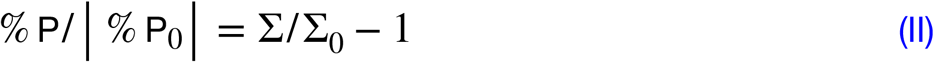

The results are shown in the following graph:

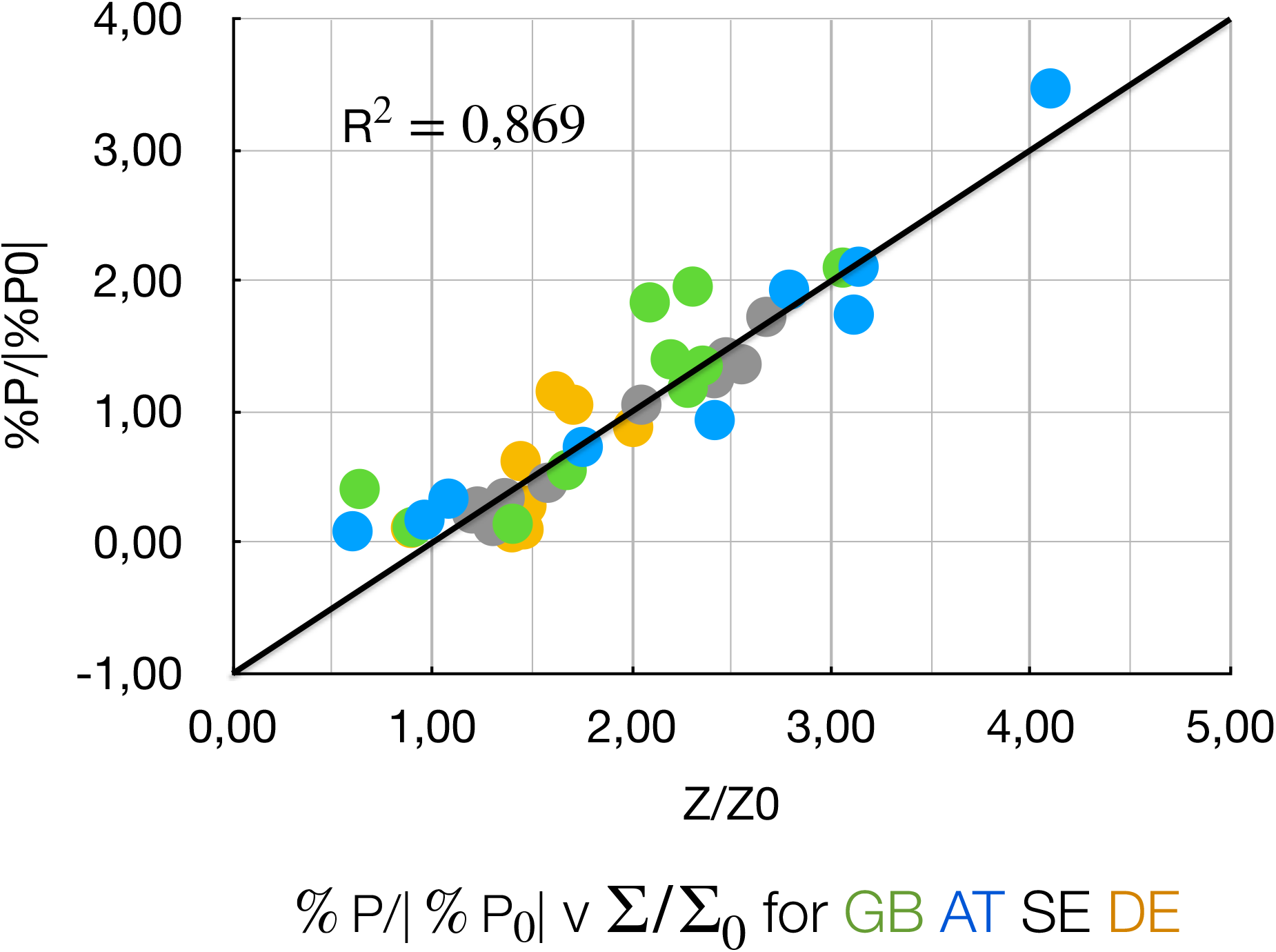

Moreover, the same equation holds for PCR positivity in London, using data from Yeadon et al [6], and in the other English regions, using data from Fenton et al [5], for the period of September - December 2020. After scaling in the same way, the following graph results:

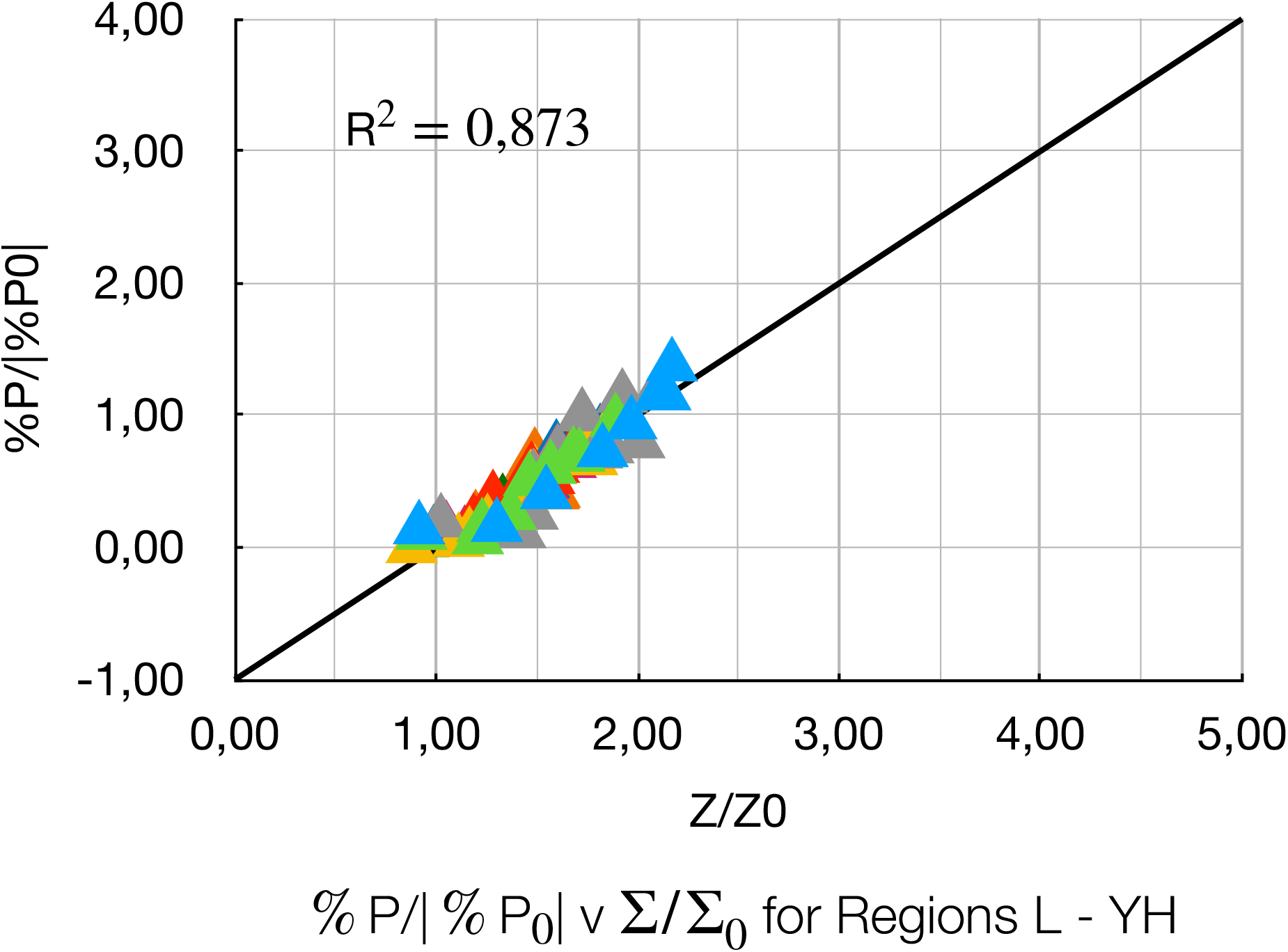

Fenton et al also provide data for 8 randomly selected local health authorities: Amber Valley, Ashford, Newham, North Tyneside, Portsmouth, Redbridge, Richmond on Thames, and Salford in England. These data can be treated analogously to yield the following graph:

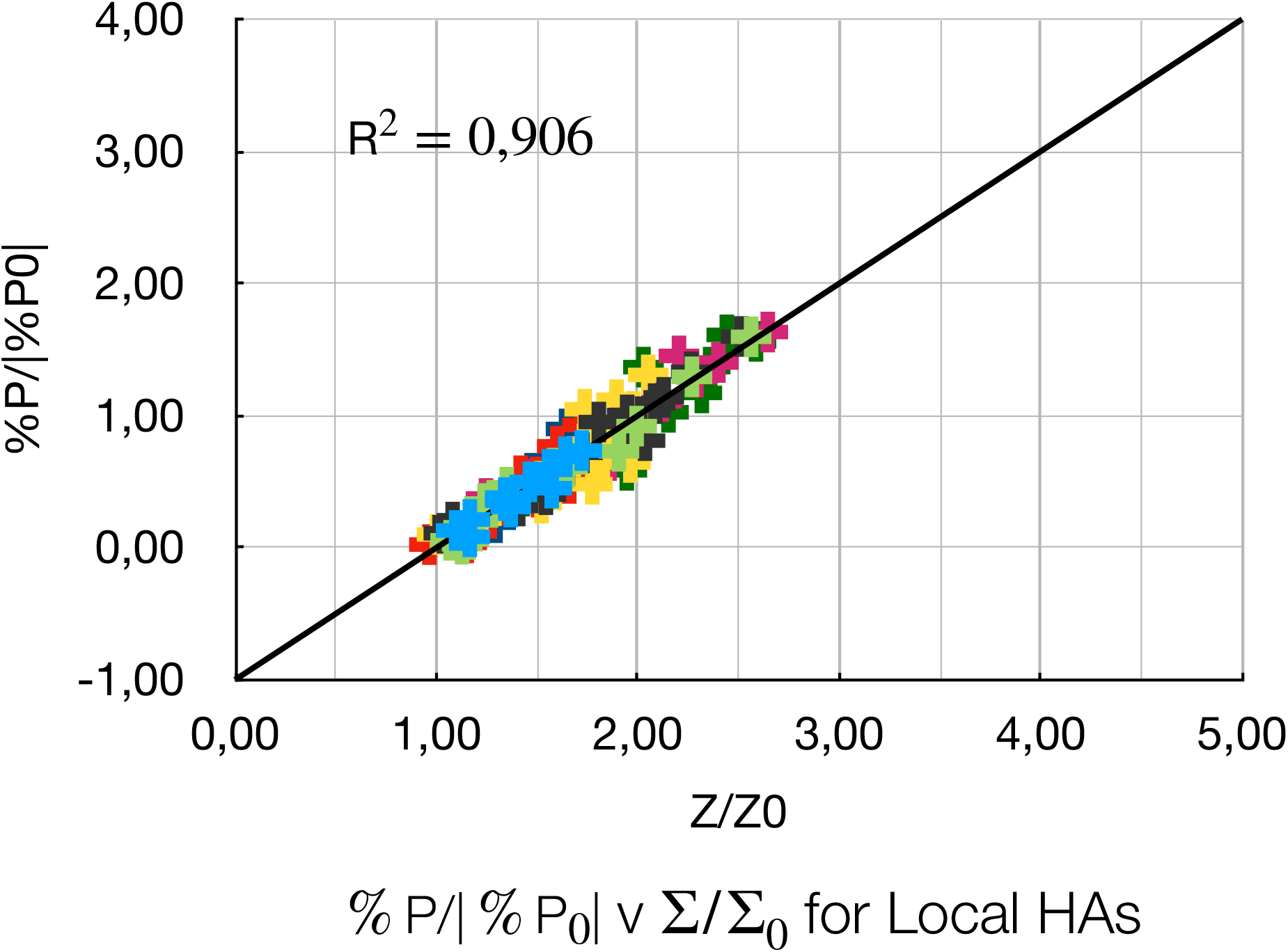

Finally, the graphs may be combined in a single plot to yield:

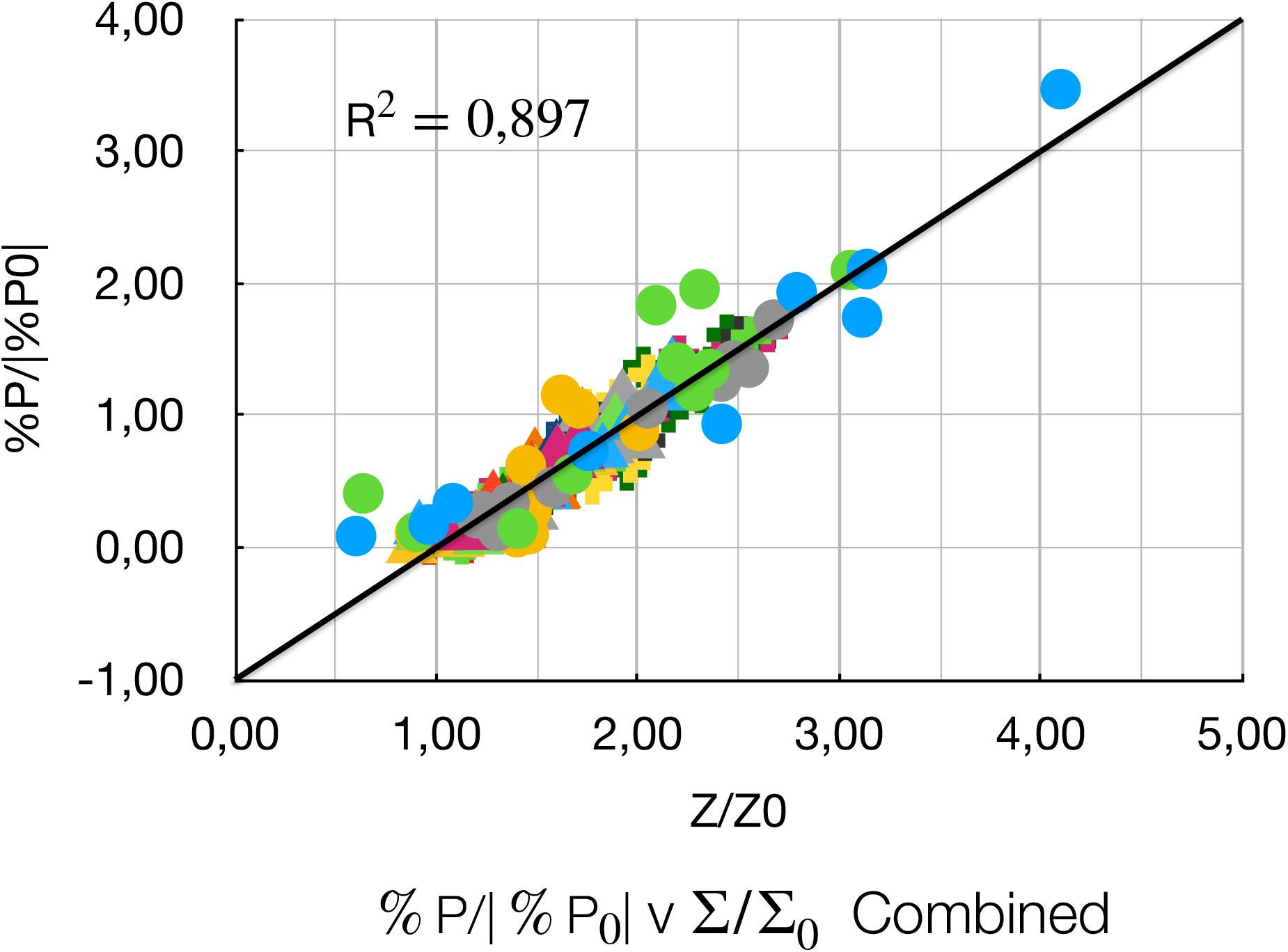

The plot comprises 21 sets of data, from 4 nations, 9 regions and 8 localities, and over 200 data points, with a co-efficient of determination, R^2^= 0,897. Thus the same scaling law applies, independent of granularity, across national, regional and local boundaries. Particularly striking, at least in the national figures, is the fact that the data appear to cluster around integral or half integral values of Σ/ Σ_0_, viz. 1/2, 1, 3/2, 2, 5/2, 3, 4.

## III. Dorfman Pooling: A Possible Explanation for Scaling

As shown in Annex II, Equation (II) follows on the assumptions of low prevalence and that the false positive rate, FPR, is a linear function of the number of tests, Σ.

One possible explanation for this particular form of the equation could be the use of Dorfman pooling [7] of individual samples in order to achieve high throughput, up to 600K tests a day. It is to be noted that the CDC PCR instruction manual expressly licenses pooling [8]. The Roche fact sheet on PCR testing for SARS-COV2 recommends pooling [9] and the NHS issued a standard operating procedure (SOP) for pooling of asymptomatic SARS-COV2 samples for PCR testing [10]. There have also been a flurry of papers on advanced pooling strategies [11-13]. In principle, if a pool tests positive, the individual samples are supposed to be retested to identify the source but under pressure to achieve high throughput, it is easy to see how this second stage might well be left out, or the results not recorded [14].

Dorfman pooling only offers an advantage at low prevalence, where false positives (FPs) predominate. So if a FP, at a rate of 0.8%, say, were to enter a pool of ten, the positivity would be amplified up to 8%. On this basis, the Austrian line in the first graph is steeper than the GB line, because with fewer facilities, larger pools are required to achieve the same number of tests. Thus | % P_0_| is a measure of the intrinsic false positive rate, FPR_0_, and Σ_0_, the intrinsic testing capacity. The ratio of the x-value to the x-intercept, Σ/ Σ_0_ gives a measure of the relative pool size. Evidence of this, as noted above with regard to the national figures, is that the data appear to cluster around integral or half integral values of Σ/ Σ_0_, viz. 1/2, 1, 3/2, 2, 5/2, 3, 4.

Nevertheless it has been argued several times [15] that only people with symptoms are tested. Consequently, as the prevalence of COVID-19 increases, the number of TPs would increase, along with the number of tests, and hence there is bound to be a correlation between the positivity and the number of tests. On this argument, FPs are a red herring.

A more sophisticated version of the argument [16] holds that testing is restricted to people who have symptoms that are compatible with COVID-19 but many of these symptoms turn out to be symptoms of something else; there is always a fair amount of respiratory and similar disease around, and it tends to be more prevalent in the winter. The number of tests carried out therefore increases over the autumn. If COVID-19 cases increase more strongly, but from a very small percentage of total respiratory disease, then there will automatically be a strong correlation between testing numbers and positivity.

In fact, according to Aukema et al [17], who use a Bayesian model to simulate PCR data for a variety of combinations of prevalence, specificity and sensitivity, there are always several possible *ad hoc* scenarios, which account for the daily figures: At the two extremes, low prevalence, high FPs scenarios and high prevalence, low FP scenarios.

In such a Bayesian model, the positivity, %P, is given by:

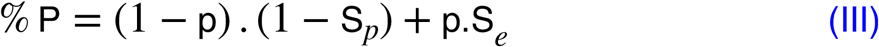

where p is the prevalence, S_*p*_ is the specificity and S_*e*_ is the sensitivity. The first term on the right corresponds to the false positive rate, FPR; the second term to the true positive,TPR.

If the number of tests, Σ, correlates with the prevalence, p, i.e.:

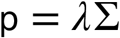

then:

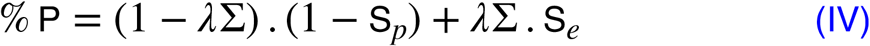

This means as p and Σ increase, P will tend to *λ*Σ.S_*e*_, i.e. a graph of % P against Σ will indeed be a straight line. Yet, as indicated in the following sketch:

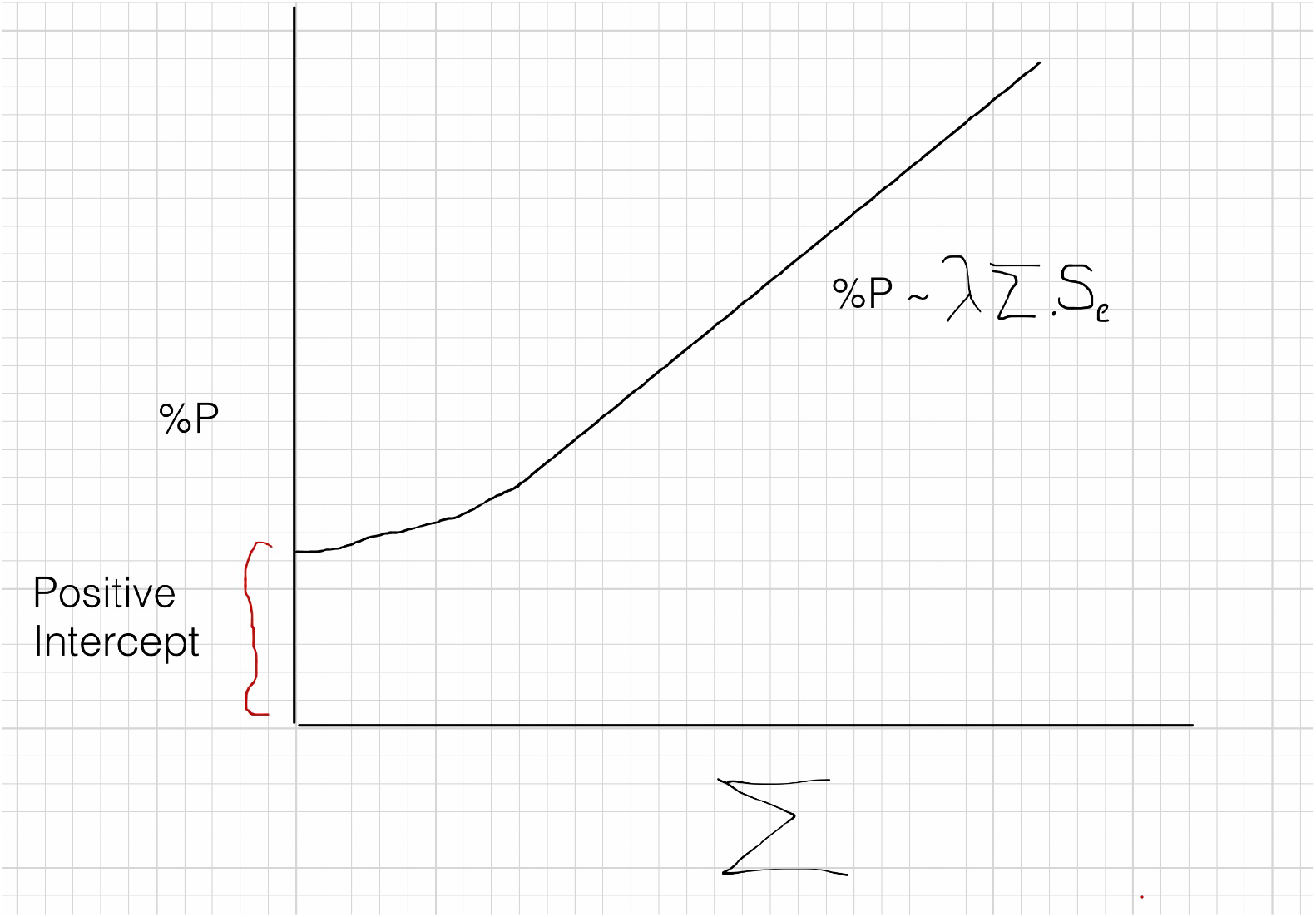

these scenarios would give a graph with a positive intercept, whereas the actual data clearly show a negative intercept, independent of any assumptions about FPs etc. Nor is there any indication how these scenarios lead to scaling across countries, regions and localities. Consequently, such high prevalence, low FP scenarios are not consistent with the data.

This leaves pooling as the most plausible explanation. Of course, it may not be official Dorfman pooling, since a consistent carry over of a positive sample into adjacent ones would yield a similar effect.

Nonetheless, whatever the explanation, some 90% of the variance in the data is accounted for by a systematic effect, which operates over national, regional and local boundaries. In the next section, the implications this has for real cases are explored, using the UK data by way of example.

## IV. How Many Real Cases?

The scaling law can now be employed to calculate the systematic or FP contribution to the daily cases. First the positivity accounted for by the regression is calculated via Equation (I) using the daily number of tests, Σ, and the parameters in Table (I). This is then multiplied by the number of tests and subtracted from actual number of positive tests, or cases, taken from OWID and represented by the blue line, a 7 day rolling average, in the graph below. The result is shown by the green line in the graph:

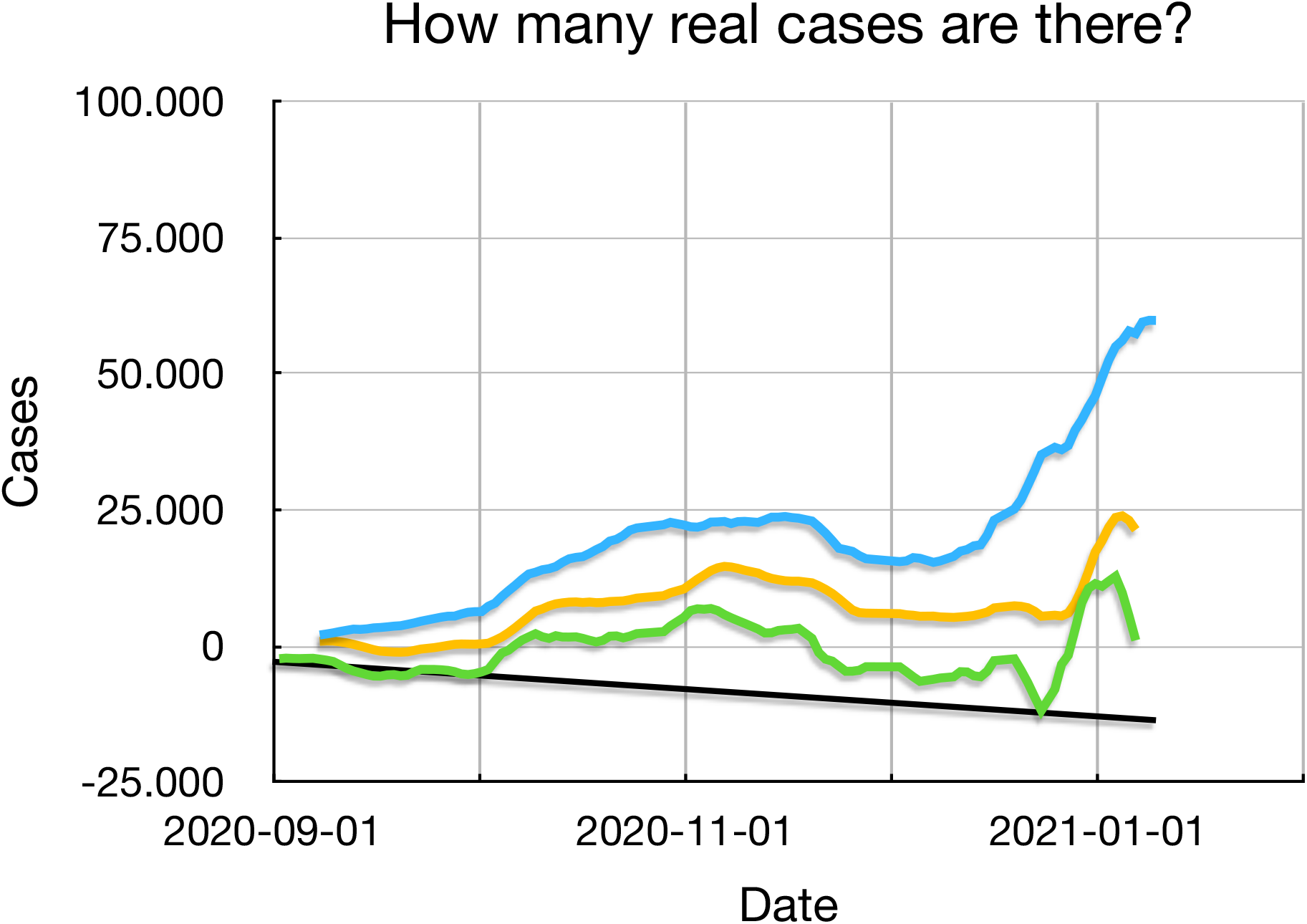

The negative going baseline, in black, deserves comment. It arises for two reasons. First, the presence of TPs will skew the regression line steeper, leading to an overestimation of the FPs by the regression. Secondly, it follows from Equation (III), that the presence of TPs suppresses the actual number of FPs, exacerbating the overestimation further. Nevertheless, the baseline can be fitted accurately to a straight line (R^2^ = 0,955) and the data corrected for the baseline to yield the yellow line, representing corrected or real cases, again as a 7 day rolling average. The curve peaked before the end of the year, with a maximum corresponding to about 45% of that of the blue curve.

The results can be compared with other data in order to corroborate the analysis. The blue line in the graph below shows daily new cases reported by the ZOE Covid symptoms tracking app, developed by Spector et al [18], over the same period. The data are for London normalised to 100K of population. The green line represents real cases from the present analysis, also normalised to 100K of population, and time shifted back one step, or 10 days. As can be seen from the subsequent correlation graph, the two sets of data are highly correlated, with R^2^=0,787. That the real cases peak at about 38 per 100K of population compared to 213 for the ZOE data is understandable, since not all those reporting symptoms via ZOE will actually be suffering from COVID-19. Similarly, it would be natural to report symptoms before seeking a test, hence the time delay.

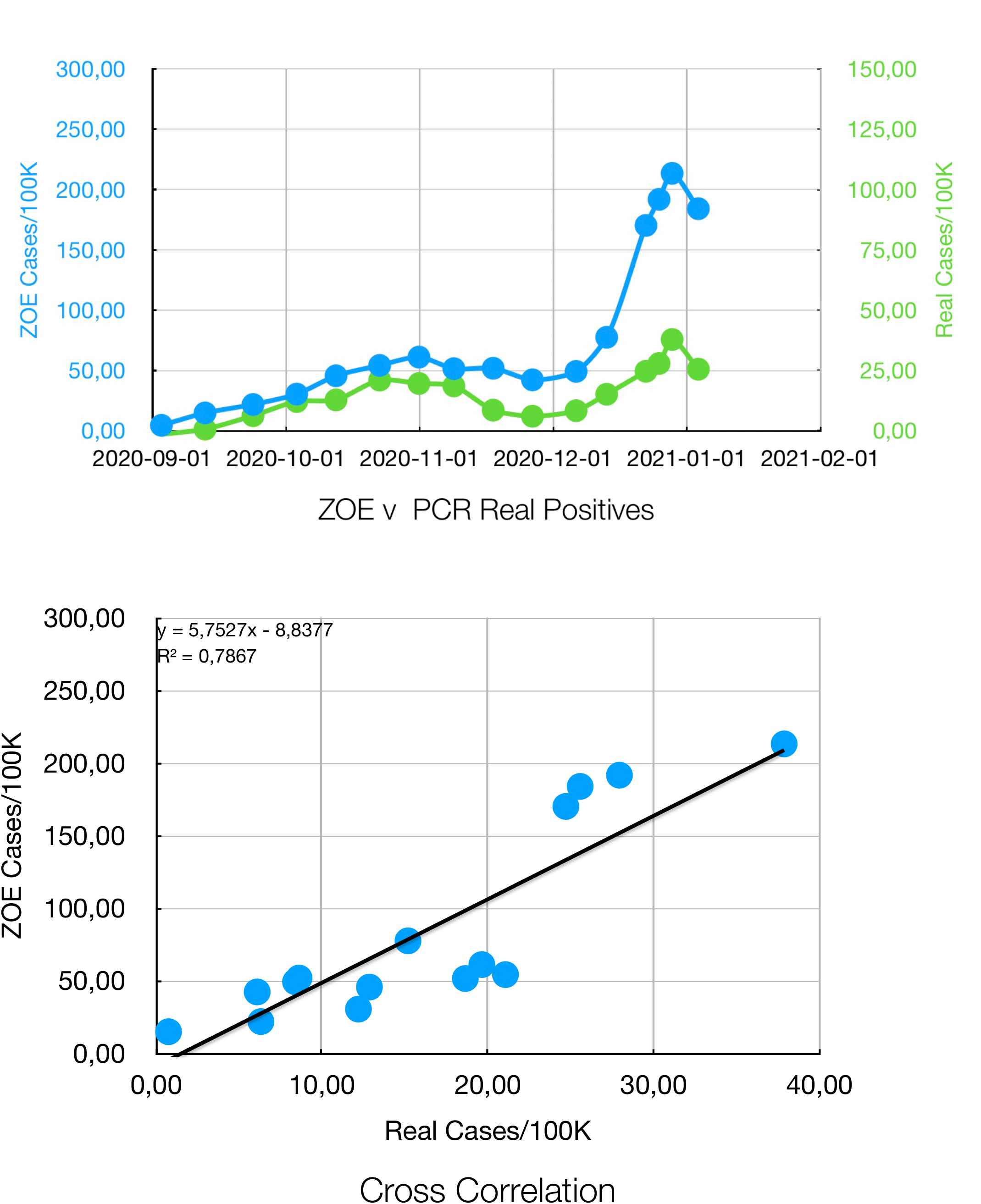

Further corroboration comes from comparing the results from the present analysis with excess mortality as estimated by PHE for the same period [19]. The blue line in the graph below represents real cases, again normalised to 100K of population and time shifted this time forward by 28 days to achieve maximum correlation, although reasonable correlation occurs for time shifts between 21-28 days. The continuous black line shows excess mortality for England normalised to 100K of population. The dashed line gives that for London. Again, the subsequent correlation graph confirms that real cases from the present analysis and excess mortality for England are highly correlated, with R^2^ = 0,833. The correlation with the London data was less good (R^2^ = 0,665).

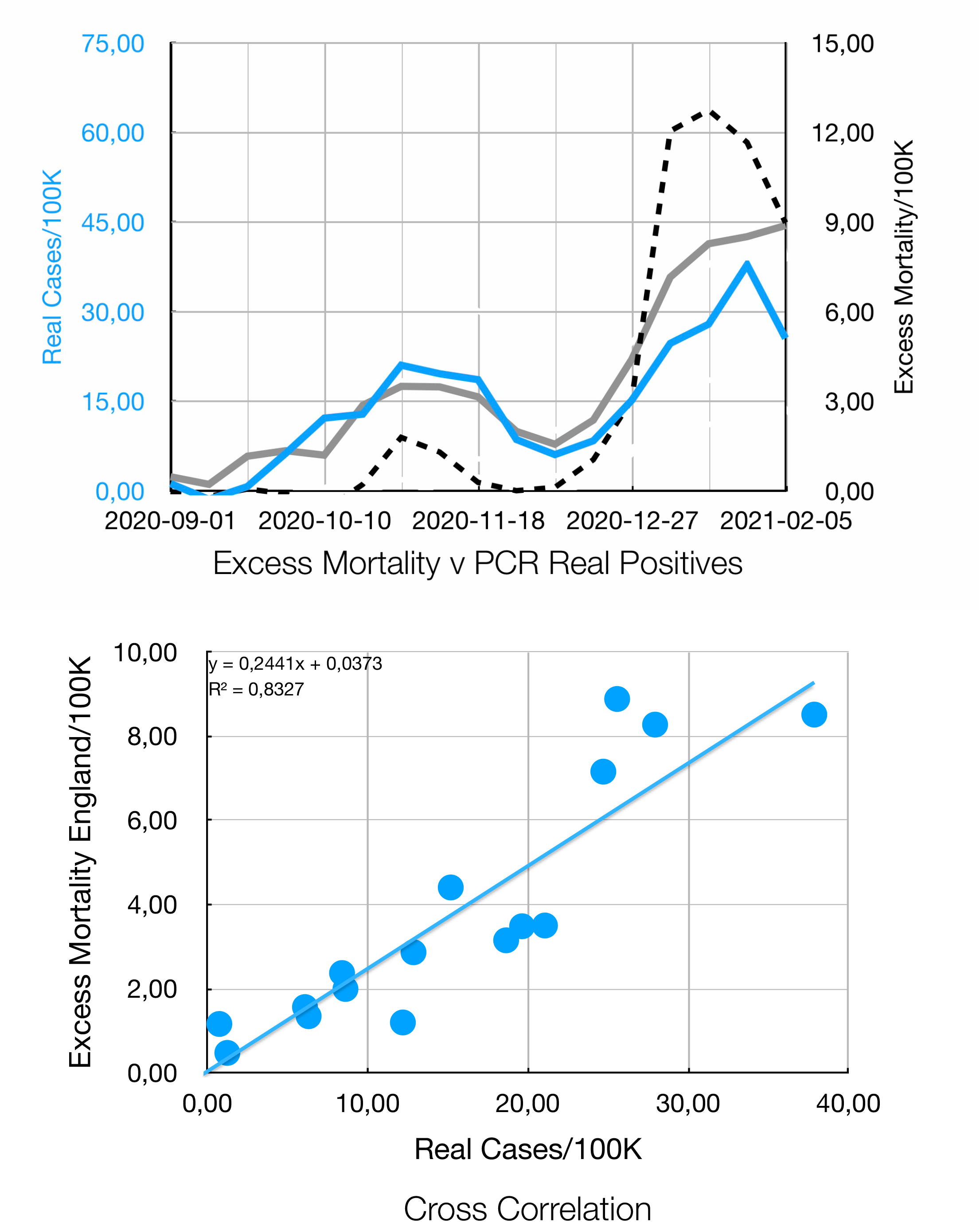

## V. False Positives

Having validated the analysis in this way, the real cases can be subtracted from the daily number of positives to yield the daily FPs. In the graph below, the middle panel shows the total daily positives in red and the daily FPs in blue, both as a 7 day rolling average. The bottom panel gives the percentage of FPs and the top panel shows the cumulative total of FPs as a function of date. This yields a figure of 1,4 million by the start of 2021. Other estimates arrive at a similar total [20].

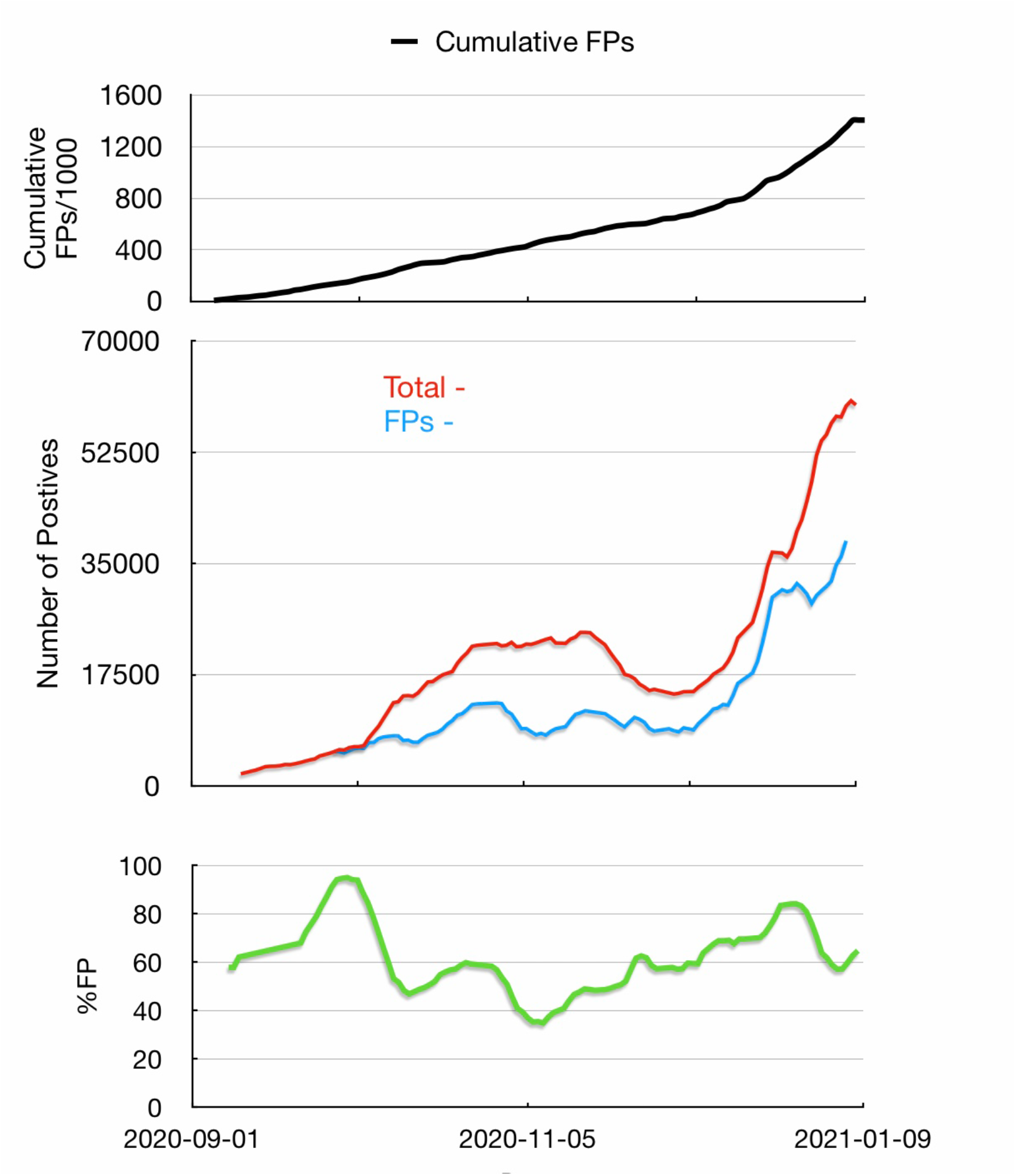

## VI. Conclusions

PCR positivity in the second wave of COVID-19 has been found to obey a scaling law, which applies across national, regional and local boundaries. The scaling law can be used to extract real cases from total daily positives. The results correlate strongly with data from the ZOE Covid tracking app and PHE data for excess mortality. By subtracting the results from the daily total the cumulative total of FPs can be estimated as 1,4 M by the start of 2021.

## Data Availability

All data produced in the present study are available upon reasonable request from the author

## Acknowledgements

The author appreciates discussions with Dr Steven Hammer and Dr Tanya Klymenko.

## Annex I - Data and Methods

Positivity data and case numbers for GB, AT, DE, SE were obtained by manually sampling plots from OWID via the cursor at approximately ten day intervals and copying to a Numbers spreadsheet (*PCR Positivity*) for analysis. Graphs for regional and local positivity and case numbers from Yeadon et al [6] and Fenton et al [5] were digitised manually via web digitiser https://apps.automeris.io/wpd/ and copied to Numbers spreadsheets (*Regional PCR positivity, Local PCR positivity*). The scaled data were transferred to spreadsheet *PCR positivity*.

To determine real cases, the full set of positivity data was downloaded from OWID and copied to a Numbers spread sheet (*owid-covid-data*). The scaling law was used to determine FPs, which were then subtracted from the total positives in the spreadsheet. The data for the baseline were obtained by digitising a plot of real cases and analysed in Numbers spreadsheet *Baseline Dataset*. The linear fit to the baseline was then used to correct the real cases in the *owid-covid-data* spreadsheet, as shown in figure 7 above.

For comparison purposes, ZOE data were obtained by manually digitising the plot for London obtained from Twitter using the web digitiser and copied to Numbers spreadsheet *ZOE Data*. The corrected cases from *owid-covid-data* were sampled at the corresponding dates and copied across to *ZOE data*. The Christmas recordings for 21.12, 23.12 and 26.12.2020 contained a glitch which was resolved by summing the individual values and posting to 26.12.2020. Alternatively, the 7 day rolling average could be used instead. The results are plotted in figures 8 and 9.

Excess mortality data, based on PHE estimates for September 2020 - February 2021, were taken across to the *ZOE data* spreadsheet from Davies, SHU Final Year Dissertation [19] and normalised to 100K of population. The results are plotted against real cases in figures 10 and 11.

Finally, collective data from *owid-covid-data* spreadsheet were transferred to Numbers spreadsheet *PCR Cases v Ct* to allow comparison with ONS Ct data where the cumulative total of FPs was calculated and plotted in figure 12.

Exemplary data sets from the the various spreadsheets are attached in the Supplementary Information. The complete spreadsheets can be downloaded from iCloud on request to the author.

## Annex II - Derivation of the Scaling Equation

By definition,

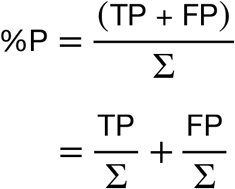

where %P is the positivity, TP, true positives, FP, false positives and Σ, the number of tests.

Suppose

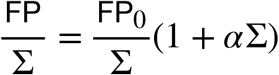

Then

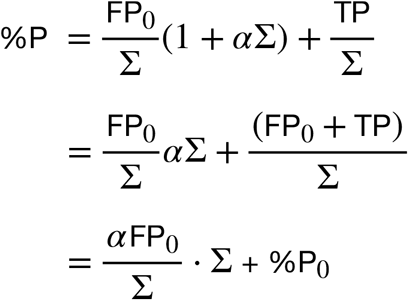

If TP ∼ 0, then 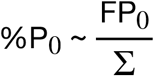 and

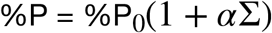

When Σ= 0, %P = %P_0_ = FPR_0_, the intrinsic false positive rate.

When 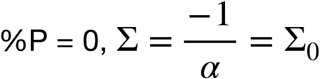, the intrinsic testing capacity.

So

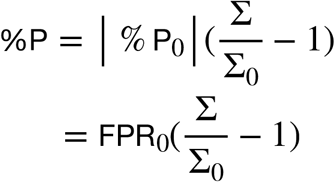

whereby, if n_0_ is the initial pool size, the false positive rate for an individual sample is

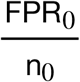

Dividing both sides by | %P_0_ | yields:

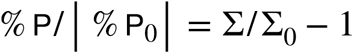

## Footnotes

1 See Annex I for details of data analysis.

